# Diffuse optical tomography system for acute traumatic brain injury in the intensive care unit: a prospective study on healthy volunteers

**DOI:** 10.1101/2024.10.09.24315200

**Authors:** Mario Forcione, Antonio Maria Chiarelli, David Perpetuini, Guy A. Perkins, Andrew R. Stevens, David J. Davies, Antonio Belli

## Abstract

**Significance:** Current systems for diffuse optical tomography (DOT) are unsuitable for clinical applications on acute traumatic brain injury (TBI) patients while in the intensive care unit (ICU).

**Aim:** To develop and test a method for DOT recordings suitable for TBI patients in the ICU. This method is based on measurements and co-registration using 3-D optical scans, and the acquisition of optical data using a custom-made helmet which would enable a multimodal (invasive and non-invasive) neuromonitoring.

**Approach:** Probe displacements compared to electromagnetic digitization co-registrations were assessed. The capacity to isolate and monitor, using functional near-infrared spectroscopy (fNIRS), the optical signal in the intracranial (ICT) and extracranial tissues (ECT) was tested on 23 healthy volunteers. Participants were scanned with a frequency-domain NIRS device (690 and 830 nm) during 5 Valsalva maneuvers (VM) in a simulated ICU environment.

**Results:** The results showed an average error in probe displacement of 5.5 mm, a sufficient capacity to isolate oxyhemoglobin O2Hb (p=6.4·10^-6^) and total hemoglobin HbT (p=2.8·10^-5^) in the ICT from the ECT, and to follow the changes of hemoglobin in the ICT during the VM (O2Hb, p=9.2·10^-4^; HbT, p=1.0·10^-3^).

**Conclusions:** The developed approach appears to be suitable for use on TBI patients in the ICU.

## 1 Introduction

Traumatic brain injury (TBI) is a leading cause of injury-related death and disability, resulting in an estimated one million deaths per year and shortened life expectancy for survivors by an average of six years (1). Currently, there is no disease-modifying treatment for the primary injury sustained by the brain during trauma, and the treatment for severe TBI remains limited to the prevention of subsequent insults (e.g., intracranial hypertension, ischemia), known as secondary brain injury.

Pivotal components of established treatment strategies in TBI patients are continuous neuromonitoring and adaptation of the physiological parameters accordingly to maintain intra- cranial homeostasis.

Functional near-infrared spectroscopy (fNIRS) represents a non-invasive, continuous neuromonitoring tool for tracking changes in brain oxygenation in severe TBI patients by measuring changes of oxyhemoglobin (O2Hb) and deoxyhemoglobin (HHb) concentrations (2–4). Contrast-enhanced NIRS can also be employed to detect changes in optical density during a dye passage (e.g., indocyanine green [ICG]), allowing one to assess cerebral perfusion and blood-brain barrier (BBB) damage (5–7). So far, commercially available NIRS devices (e.g., INVOS 5100 Cerebral Oximeter; ISS Oxiplex TS) have not shown sufficient ability to detect episodes of ischemia in cases of severe TBI compared to the invasive intracranial techniques (e.g., brain partial oxygen pressure [PbtO2]) (8–10). However, the validity of a direct comparison between (i) the level of cerebral tissue saturation and its fluctuations measured by the optical device, and (ii) the absolute values and their changes measured by the PbtO2 monitor, can be impaired by the complex pathogenesis of brain trauma (11). In contrast, the addition of fNIRS and contrast-enhanced NIRS to currently used neuromonitoring techniques, including PbtO2, could potentially lead to a more precise assessment of the tissue status (5, 11). Thereby, the drive for ”non-invasive equivalence” to PbtO2 should be dropped, and instead the hitherto untested abilities of the optical technique, when included in a multimodal monitoring regime, to stratify injury severity and aid clinical decision-making, could finally be explored.

Diffuse optical tomography (DOT) is an extension of NIRS imaging that uses a distributed high-density array of NIRS probes to spatially map tissue optical properties on a co-registered structural image (e.g., computerized tomography [CT], magnetic resonance imaging [MRI]) (12, 13).

Compared to commercially available NIRS devices, DOT offers several advantages in the TBI monitoring by addressing the anatomical complexity of the TBI lesions (11). This advantage suggests that the commercially available NIRS devices used in clinical trials to date have not fully explored the capabilities of optical techniques and that DOT should be tested instead. Whereas DOT has been successfully demonstrated as a neuromonitoring technique in infants admitted to the neonatal intensive care unit (ICU), the authors are not aware of any studies which have performed DOT measurements on adult severe TBI patients in the ICU (14). One of the difficulties in performing such studies is related to the clinical environment in which the optical data should be acquired (e.g., contamination of the signal from the ambient light, lack of space at the patient’s bedside, presence of an intracranial bolt for intracranial pressure [ICP] neuromonitoring). Furthermore, the standard processes of probe digitization using an electromagnetic digitizer, or of adding skin markers to the subject’s structural imaging, would be impractical solutions to implement DOT in the clinical practice of brain trauma care (15–17): the electromagnetic signal used by common digitizers can be impaired by the presence of metal at patients’ bedsides; the optical analysis can only be performed *after* skin markers have been added. Since a CT scan is one of the first neuroimaging assessments made on TBI patients upon hospital admission, it is usually performed urgently (18). This makes it unlikely that skin markers could be positioned on TBI patients, at the locations where the optodes will be placed, prior to their first scan. Furthermore, it should be taken into consideration that the CT scan is crucial to the clinical decision process, and the presence of skin markers may increase the risk of imaging artefacts which can impair accurate radiological assessment. Therefore, a method that co-registers the optical data into a CT scan without the aid of skin markers, or electromagnetic digitization, is needed to make DOT analysis on TBI patients possible from the point of admission.

We devised a system to perform DOT recordings on acute TBI patients from the point of admission into ICU, comprised of a custom-made helmet for optical data acquisition in a multimodal neuromonitoring (e.g., intracranial probes), and a co-registration of the optical data on patients’ structural images acquired for clinical assessment (e.g., CT scans). On healthy volunteers, we tested: (i) the accuracy of the novel co-registration compared to one based on electromagnetic digitization; and (ii) the capacity to accurately detect changes in levels of Hb in the ICT and separate signals from ICT and ECT, during VM.

## 2 Materials and Methods

### 2.1 Participants

23 healthy volunteers (17 males, ages ranging between 20 and 42 years old with a mean of 28 years) were recruited into the RECOS study (Repetitive Concussion in Sport) (IRAS ID: 216703; Ref. REC: 17/EE/0275) performed at Centre for Human Brain Health, University of Birmingham, UK. They had different hair colours and hair densities.

Participants took part in this study after providing their written, informed consent. The study was conformed to the Declaration of Helsinki and was approved by the East of England-Essex Research Ethics Committee on 22 September 2017.

### 2.1 Equipment

#### 2.1.1 Optical helmet

A custom-built optical helmet was adapted from the description of Tan et al. to make it suitable for optical recordings on TBI patients in the ICU (Figure 1) (19).

**Figure 1.**
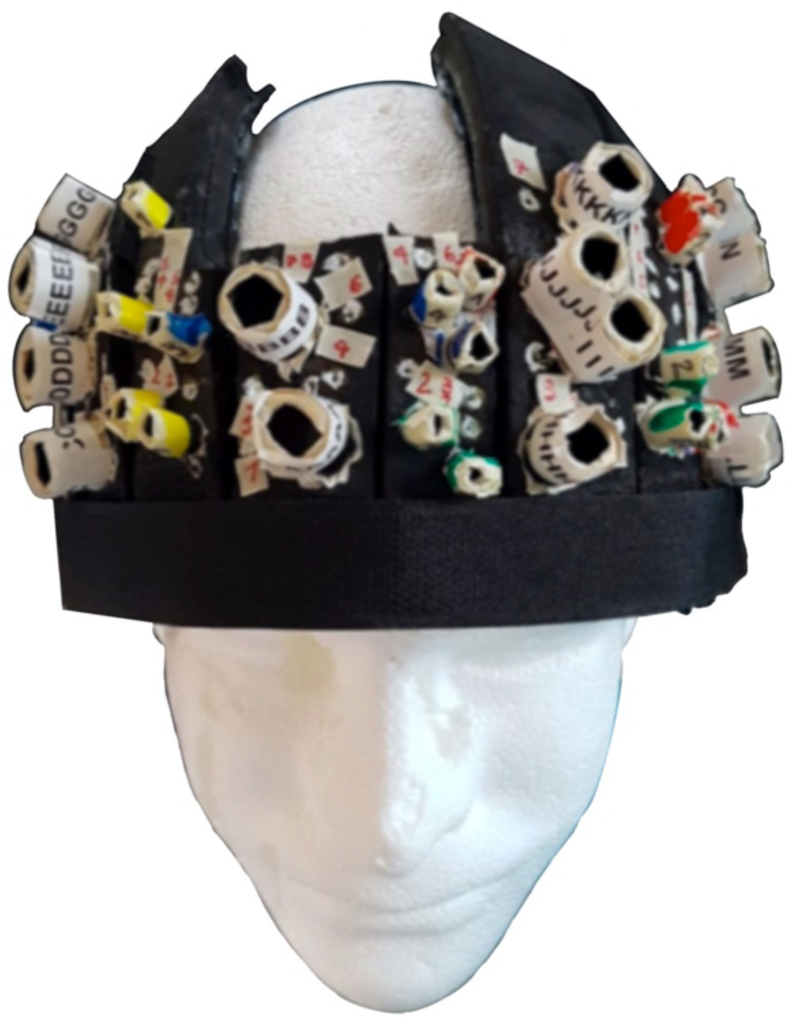
Picture of the helmet. The design features an open area at the top of the skull to avoid interference with invasive neuromonitoring devices.

The helmet comprised multiple adjustable segments. It could be adjusted to fit a variety of head shapes and sizes, including post-surgery (e.g., decompressive craniectomies; external ventricular drains (EVD)). It was vital that the helmet design maintained patient comfort and safety (e.g., no increase in ICP), whilst also being rigid enough to keep the probes still during the recording. The helmet is designed to be affixed to the patient using disposable Velcro straps or medical bandages under the chin. To reduce the risk of infection and to grant access for intracranial monitoring and treatment, no slices were positioned in the vicinity of the Kocher’s point, where the ICP bolt and the external ventricular drain (EVD) are usually positioned (18).

The helmet comprised multiple black layers. The thicker part was made of two layers of black foam (The Michaels Companies, Las Colinas, Irving, Texas, USA) to shield the probes from ambient light as well as to keep the helmet lightweight. These two layers were glued together while held in a curved shape, to model the helmet frame. The internal lining of the helmet was made of a thin layer of polyurethane rubber, RenCast (Freeman manufacturing and Supply Company, Avon, Ohio, USA), which came into contact with the scalp, along with the probes. Polyurethane rubber has been used in previous studies on TBI patients in the ICU and, in our experience, it increases the helmet’s integrity whilst also being suitable for clinical environments, as it is non-porous, soft and easy to clean (8, 20).

The layout for optical recording covered only the frontal lobes to facilitate the positioning of the probes on TBI patients who have limited head movement due to being unconscious in a supine position with their head being tilted-up, as per ICP control care, potentially wearing a hard cervical collar, and being intubated or with a tracheostomy (18) (Figure 2, A).

**Figure 2.**
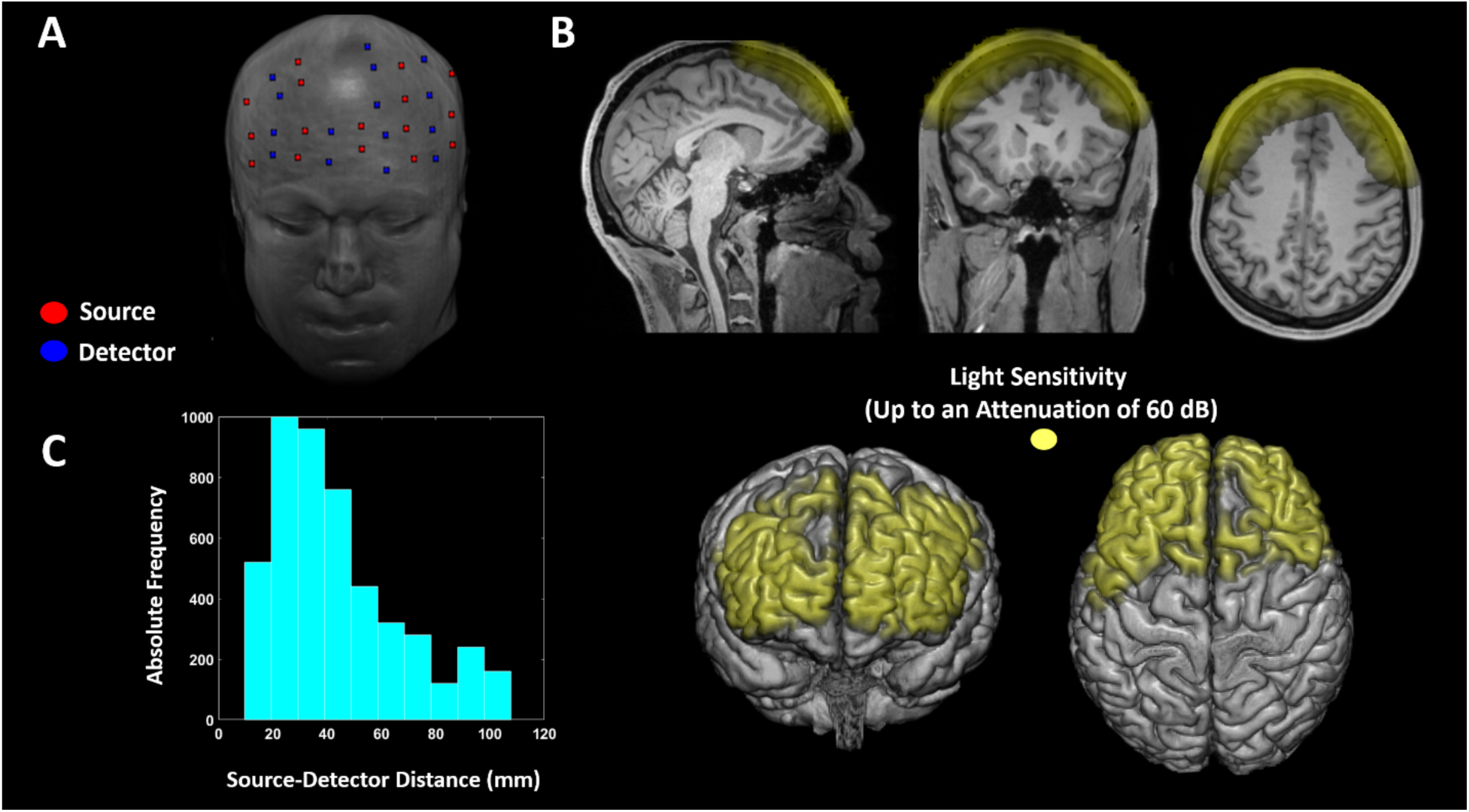
A: Illustration of the optical probes’ positions on a structural MRI. The sources’ positions are illustrated as red dots and the detectors’ positions as blue dots. B: Average light sensitivity of the probe configuration used, computed with a FEM approach (21). The light sensitivity is displayed up to an attenuation of 60 dB (1000 times), and it is overlaid on an MRI of a representative participant. C: Absolute frequency of the SD distances of the channels across all subjects.

Before making holes to house the probes, a “3-space” FastTrak 3-D digitizer (Polhemus, Colchester, Vermont, USA) was used to electromagnetically digitize the helmet surface using the nasion and the external acoustic meatuses (EAM) of an artificial head as fiducial points. Based on the obtained digitization, a distribution of probes was selected to scan the broadest possible area with the maximal number of SD distances between 25 and 35 mm, so as to maximise sensitivity to the ICT (12) (Figure 2, C). A custom-made MATLAB graphical user interface, near-infrared optode montage automated design (NOMAD) (https://github.com/kylemath/nomad), developed by Mathewson et al., was used to evaluate the acceptability of a plotted configuration of SD locations on the helmet, by ensuring the illumination of the frontal lobes of both hemispheres whilst avoiding cross-talk between SD pairs in a time-multiplexing cycle of light-emission (22). The average light sensitivity of the probe configuration used was computed with a FEM approach (Figure 2, B) (21). Below 60 mm distances, the probes were deemed to be usable for fNIRS-DOT analysis based on the data recorded during the VM. Holes to position the probes were punched only where the NOMAD analysis recommended. The limited number of holes, compared to the model described by Tan et al., was necessary to reduce the amount of ambient light that reaches the probes, by ensuring that no hole went unutilized, as well as to allow data recording in a supine position (19). The large diameter of the holes where the detectors were inserted allowed for the removal of any hair between the detector and the skin, in order to maximise photon harvesting from the scalp (23, 24). A plastic cone was inserted inside the hole along with each detector to mechanically secure them to the helmet, and to reduce the signal contamination by ambient light (24). The holes were labelled and color-coded to facilitate the positioning of the probes. Small white dots, that could be easily seen on an optical 3-D scanner, and whose small size still maintained a high level of pinpoint accuracy, were drawn on the helmet in the regions that were to be exposed in a supine participant.

#### 2.1.1 Data acquisition

##### 2.1.1.1 Digitization

Participants were scanned with an optical 3-D scanner (Artec Leo, Artec 3D, Luxemburg, European Union) while wearing the helmet and sitting on a chair. The quality of signal acquisition was confirmed by a visual inspection on the 3-D scanner screen. On a 3-D image, the *x-y-z* coordinates of the fiducials (i.e. nasion, EAM) and those of the white dots were measured using complementary software (Artec Scanner, Artec 3D, Luxemburg, European Union) (Figure 3, A).

**Figure 3.**
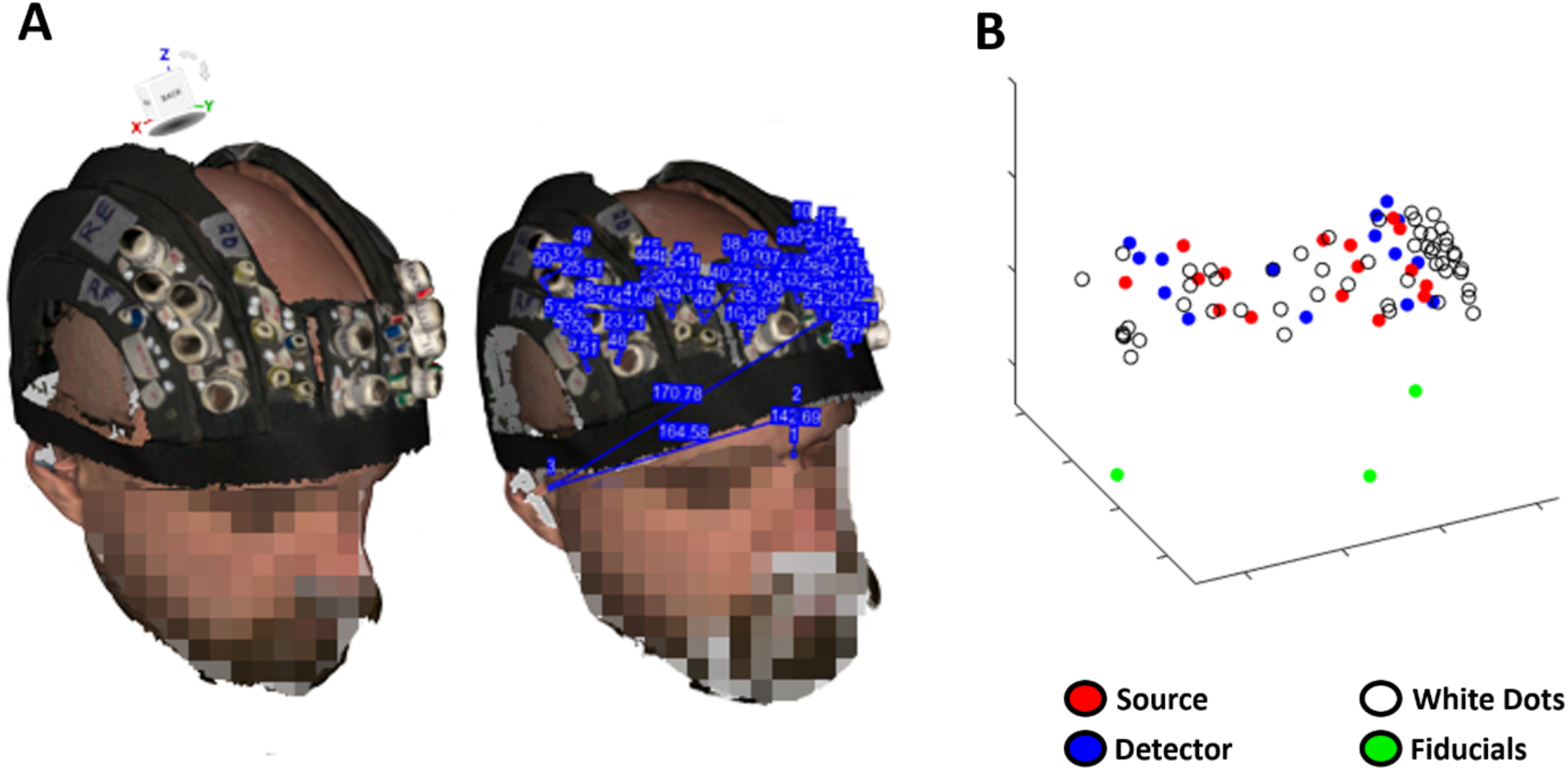
A: Picture from Artec Leo reconstructed using Artec Scanner and measurement of the 3-D coordinates of the nasion, EAM, and white dots. B: example electromagnetic digitization of fiducials (nasion and eam), white dots and optodes with FastTrak.

The fiducials, the holes for the probes, and the white dots were also digitized using the FastTrak 3-D digitizer (Figure 3, B). The nasion and the EAM were chosen as fiducials because, according to our research experience, they can be identified relatively easily on an anatomical scan without the use of skin markers.

##### 2.1.1.1 Optical recordings

Optical data were recorded with a FD-NIRS system (Imagent; ISS Inc., Champaign, IL). Data were collected from 15 detectors, each measuring light emitted by eight of the 16 total source locations (120 dual-wavelength channels). Laser diodes delivered light, modulated at 110 MHz, for each source location at 690 and 830 nm wavelength (max power: 10 mW, mean power: 1 mW). The different diodes were switched on in a time-multiplexing manner. The light from the diodes was transmitted to the scalp using thin optical fibres (diameter = 400 µm), whereas the back-scattered light from the head was collected using detector fibre bundles (diameter = 3 mm) connected to photomultiplier tubes (PMTs). Based on heterodyne detection, signal digitization, and fast Fourier Transform, the system generated temporal modulations in the emitted light DC (average), AC (amplitude), and phase. Optical data were sampled at 39.0625 Hz.

Coupling efficiency between the optodes and the skin, as well as signal noise, were monitored before the beginning of each scan using a complementary software (BOXY, ISS Inc., Champaign, IL) and adjustments were made to maximise the signal quality.

##### 2.1.1.1 Valsalva manoeuvre

The VM is a forced attempt to exhale against a closed glottis. The detailed knowledge of the expected hemodynamic changes during VM was used to validate the system presented, and so is described hereafter. Based on the changes in blood pressure, the VM can be divided into four phases (Figure 4) (25).

**Figure 4.**
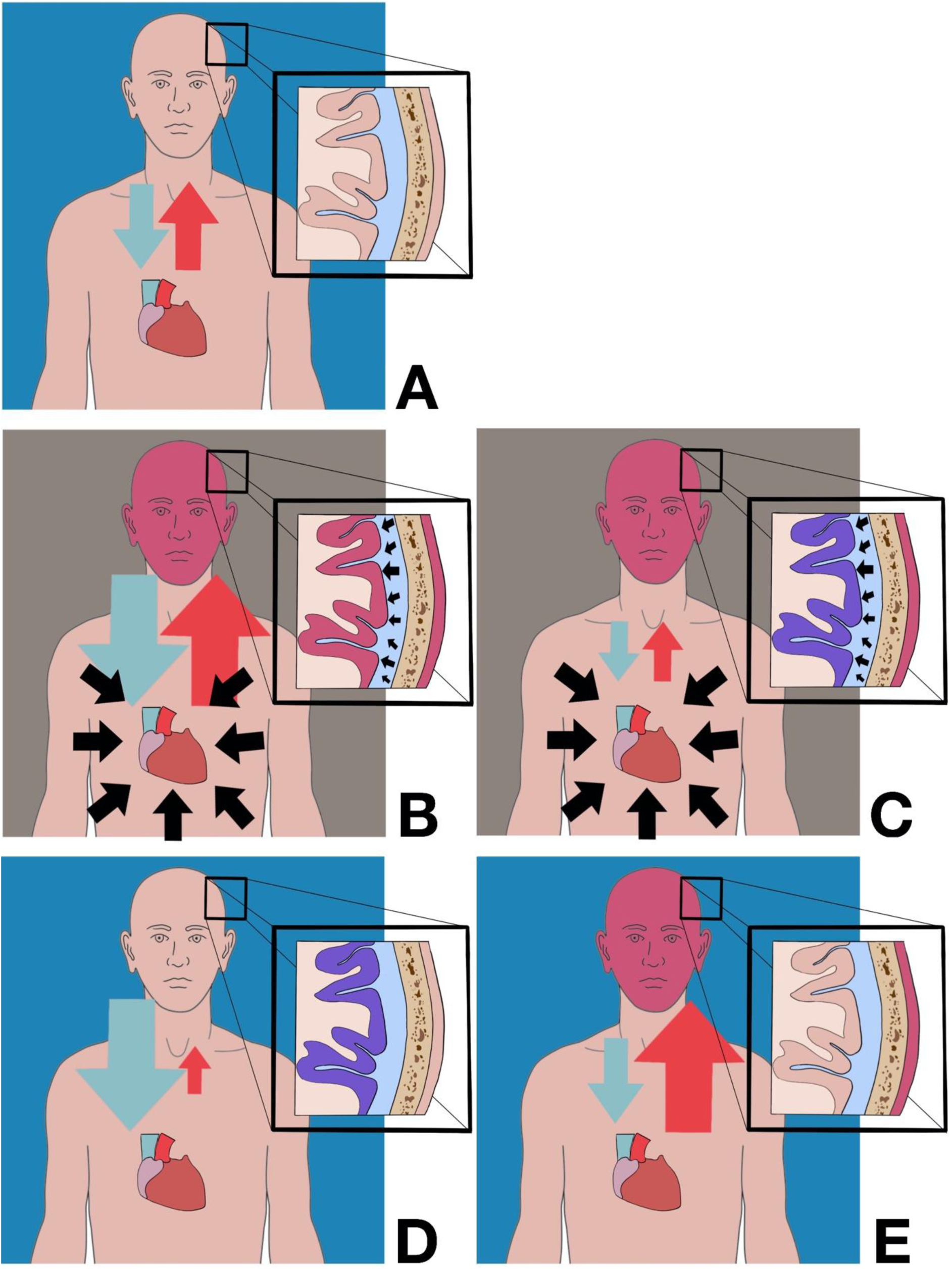
Representation of the physiological changes during the Valsalva manoeuvre’s phases. The blue and grey backgrounds represent open and closed glottis, respectively. A: standard physiological state. B: Phase I. C: Phase II. D: Phase III. E: Phase IV.

During phase I, lasting approximately 2 seconds, an increase of intrathoracic pressure, due to the forced exhalation attempt against the closed glottis, causes an increase of external pressure on the main vessels (e.g., aorta, superior vena cava) with consequent increase of the arterial and venous pressures (Figure 4 [B]) (25, 26). The increase of blood volume causes an increase in the levels of total hemoglobin (HbT) and tissue saturation in the ICT and ECT (Figure 4 [B]) (27). Due to the confinement of the brain into a non-expandable space, the increase of cerebral blood volume and the incapacity for it to drain because of the high intrathoracic pressure is immediately followed by a sharp rise in ICP (28–30). The high pressure exerted on the spinal compartment by the thorax is another contributing factor in the increase of ICP (30). This simultaneous rise in ICP serves as a protection mechanism for the brain and its blood vessels, which could otherwise be violently damaged by the sudden increase of arterial pressure (29, 30). During phase II, the reduction of end-diastolic volume, because of the reduction in venous return due to the high central venous pressure, leads to a decrease of arterial pressure and an increase of HR as a compensatory mechanism (Figure 4 [C]) (25). The reduction of blood supply, combined with the high brain metabolism, causes a progressive, non-compensated consumption of tissue O2Hb and a consequent gradual decrease of ICT saturation (Figure 4 [C]) (27, 31, 32). On the other hand, a similar reduction of tissue saturation is not present in the ECT due to its low level of metabolism (Figure 4 [C]) (32). During phase III, the glottis is opened, and this is followed by a drop of intrathoracic pressure. The sudden lack of external pressure on the main vessels determines a high level of venous return and a rapid decrease of arterial blood pressure and ICP (25, 33) (Figure 4 [D]). It is worth noting that, at this stage, the level of arterial blood pressure is at its lowest point (33). The large pool of Hb is removed from the ECT by the venous return (Figure 4 [D]). In ICT, the already low tissue saturation further decreases due to the reduced arterial blood pressure (Figure 4 [D]) (27). In phase IV, there is a peak of arterial blood pressure and a subsequent increase of the O2Hb levels in the ECT and ICT (Figure 4 [E]) (25, 27).

The VM was considered a valid task to test the neuromonitoring assessment with fNIRS-DOT, due to the substantial and opposite changes of tissue saturation in ICT and ECT. These two different behaviours may give insight on the ability of the developed DOT approach to uncouple the contributions from the ICT and ECT, and so test the capacity of the optical technique to record the brain signal without interference from the ECT (34, 35). Furthermore, the VM simulates, on healthy human subjects, a common post-brain trauma scenario, characterised by a decrease of brain saturation and an increase of ICP (32, 36–39).

Participants were instructed to perform five VMs of 10 seconds each, preceded and followed by three minutes of rest. 10 seconds before each VM, participants were asked to prepare for the upcoming physical task taking long gasps as they were deemed suitable to generate intrathoracic pressure during the VM. Participants were in a semi-supine position with their heads slightly tilted by a cushion and in an illuminated environment. A clinician was observing the participants to reduce the effects of adverse events (e.g., syncope) and to ensure that the VMs were performed correctly. The VMs were monitored by measuring the expected HR increase using a pulse-oximeter, Fingertip (ChoiceMMed, Yuquan Road, Haidian District, Beijing, PR China), and by observing signs of physical effort (e.g., face redness, shakes for the strain, breath release at the end of the VM). Participants could stop the experiment at any stage.

#### 2.1.1 Data analysis

##### 2.1.1.1 Digitization

The white dots from Artec Leo were co-registered on a structural image template through fiducial alignment based on least square fitting. The optode locations were co-registered by combining the locations of the white dots and the fiducials according to Artec Leo, with the locations of the white dots and the optodes according to the FastTrak 3-D digitizer. This method was necessary to mitigate against the potential for inaccurate optode localisation and, in extreme cases, optode omission by Artec Leo, due to blurred images, large holder sizes (especially for the detectors), and the difficulty of ascertaining the probes’ depth within the helmet. On the other hand, the digitizer is capable of measuring the coordinates of both the white dots and the optodes more reliably.

The root mean square error (RMSE) between the Artec Leo image, using the method described above, and the FastTrak 3-D digitizer; and the number of white dots necessary to optimize this approach were investigated. Finally, the co-registration of the optodes on a template was used for the optical analysis.

##### 2.1.1.2 Optical data analysis

The optical DC intensity data were transformed into optical density (OD), movement corrected, and band-pass filtered between 0.01 and 0.5 Hz (zero-lag, 2^nd^ order, Butterworth digital filter) (40, 41). For each wavelength and channel, in addition to the ten seconds of the VM trial, single-trial OD was evaluated from 20 seconds prior up to 20 seconds after the VM onset. OD signal, of 50 seconds duration, was randomly chosen during each rest period to obtain a control measurement. Average OD response for each wavelength, channel and subject was computed.

For each wavelength, only channels that provided a maximum average OD change below or equal to 0.05 (∼5% signal change) were deemed as usable. On average, 78±10 and 72±11 channels for the 830 nm wavelength and the 690 nm wavelength, respectively, were used for further analysis. Furthermore, a model of light propagation within head (forward model) and an inverse procedure were performed using a structural MRI (MPRAGE sequence) as a template. The FEM applied to the diffusion equation was used to estimate the forward model (42, 43). The FEM software NIRFAST was used to model light propagation through the head and to compute Jacobian (sensitivity) matrices of DC light intensity to absorption changes induced by Hb oscillations (21, 44). Figure 2 [B] reports an example of an average Jacobian (average optical sensitivity) for a participant overlaid onto the anatomical MRI template. The average Jacobian is displayed up to an attenuation of 60 dB (1000 times) from its maximum value. “Fine” meshes (maximum tetrahedral volume 2 mm^3^) were generated for FEM using the MATLAB software iso2mesh (45). The heterogeneous head models were based on the segmentation of the anatomical MRI. Segmentation of the skull and scalp, CSF, white matter, and grey matter was performed using Statistical Parametric Mapping (SPM) functions applied to the image (46, 47). Baseline optical properties (μ_α_, μ^*s*^ and η) of the tissues at the relevant wavelength were taken from Tian et al. (48). An inverse procedure based on energy minimisation of the solution was used to convert intensity changes on individual channels to absorption changes in voxel space at each wavelength (44). The Lambert- Beer Law was inverted to evaluate O2Hb and HHb oscillations given absorption modulation at the two wavelengths employed. The extinction coefficients of the two forms of Hb at the 690 nm and 830 nm wavelengths were extracted from Zijlstra et al. (49). Using the computed MRI segmentation, the MRI was separated into ICT and ECT compartments. The ICT compartment was evaluated by summing a smoothed version of white and grey matter segmentation. The ECT compartment was obtained by subtracting a binary image of the head with the previously defined ICT compartment. O2Hb and HHb signals were evaluated for each subject by averaging the Hb oscillations obtained in the different compartments, in a region where enough light sensitivity was obtained (-60 dB of attenuation, Figure 2 [A]). Across subjects’ average responses of O2Hb, HHb, and HbT (HbT= O2Hb +HHb) and associated statistics were evaluated.

## 3 Results

### 3.1 Co-registration

Figure 5 reports the RMSE between Artec Leo and FastTrak for the available white dots (51 in total), as a function of the number of white dots used for co-registration between the two modalities.

**Figure 5:**
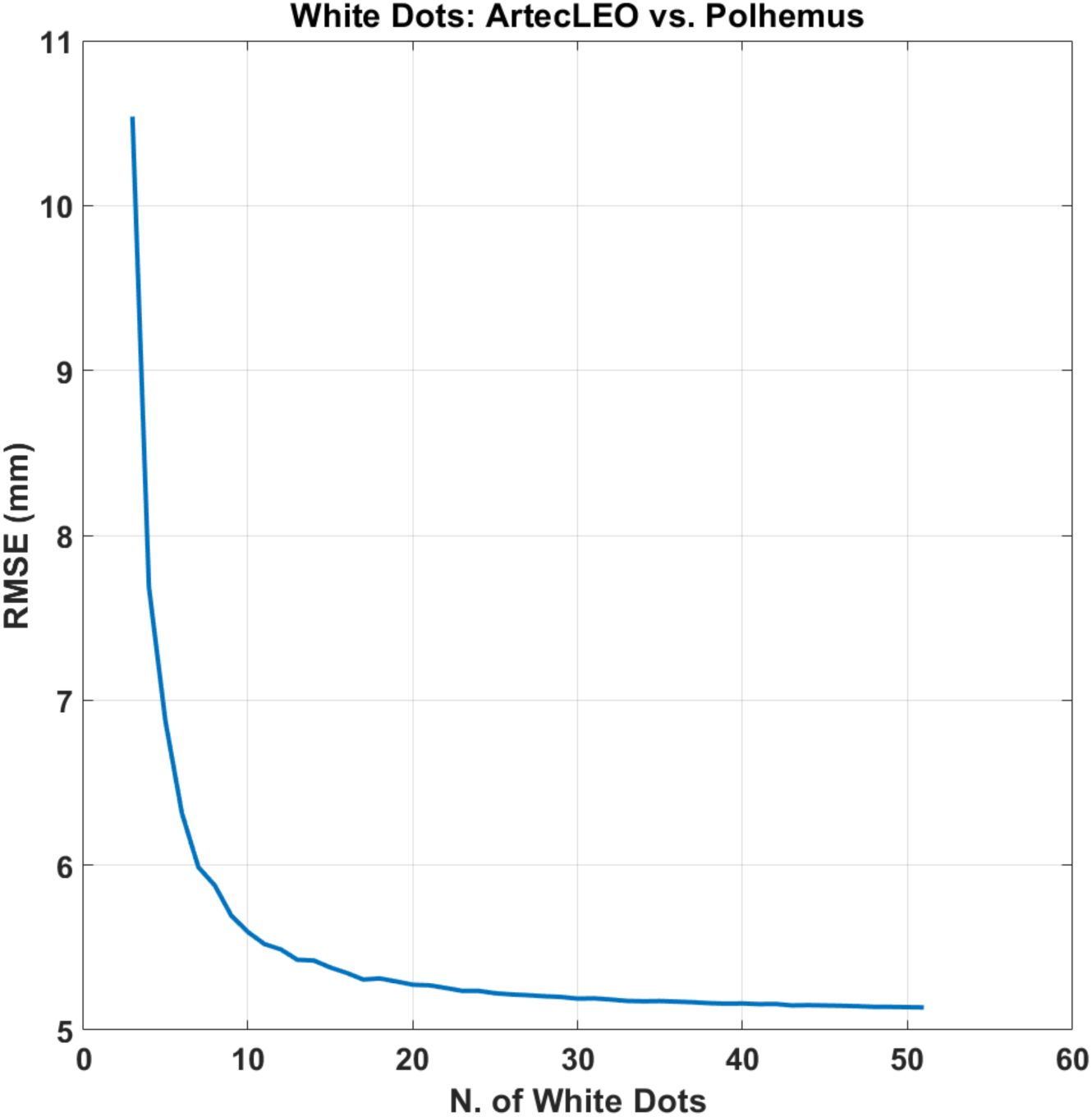
RMSE between Artec Leo and FastTrak for the available white dots (51 in total), as a function of the number of white dots used for co-registration between the two modalities. The RMSE was computed using 10,000 iterations for each number of white dots selected, where the white dots used where randomly chosen.

**Figure 6.**
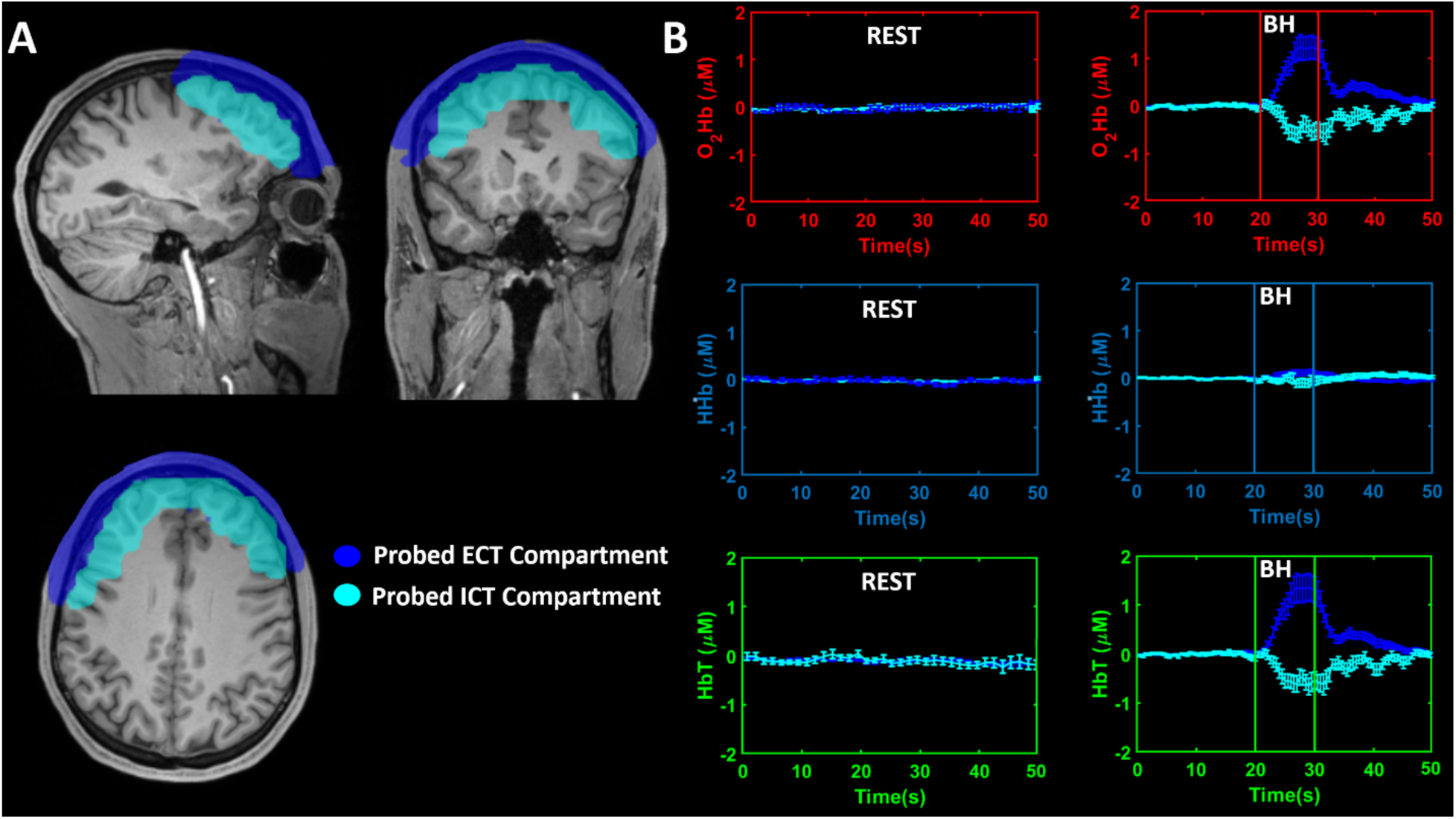
A: Computed ICT and ECT compartments that were probed by the optical array overlaid on the structural image. B: Across subjects’ average modulations, and associated standard error, of O2Hb, HHb and HbT during the rest period and the VM in the two compartments. BH: breath-holding

The RMSE reported in the figure was computed using 10,000 iterations for each number of white dots selected, where the white dots used for co-registration in each iteration were randomly chosen from all those available. The minimum number selected was 3. The plot in Figure 5 clearly shows a plateau in the error between the two modalities with a number of white dots above ten. With this number of dots, the average distance between the corresponding locations is within 5.5 mm.

### 3.1 Optical data

Five VMs were performed per participant, resulting in a mean increase in HR of 17 bpm (± 11) (t=5.17, df=21, p=3.9·10^-5^).

Figure [A] reports the ICT and ECT compartments that were probed by the optical array, overlaid onto the structural template image.

The compartments were computed based on the structural MRI employing the algorithm described. Figure [B] reports the across subjects’ average modulations, and associated standard error, of O2Hb, HHb, and HbT during the rest period and the VM in the two compartments.

A clear difference in fluctuations of levels of Hb during rest and the VM is visible. Moreover, during the VM, decoupling in the Hb responses between the ICT and ECT compartments is evident.

In both these two layers, the changes of O2Hb, HHb, and HbT are in agreement with the expected changes of tissue saturation during different VM phases. In the first few seconds of the task, the levels of O2Hb and HbT began to increase in the ECT and decrease in the ICT. Subsequently, the levels of O2Hb and HbT in the ECT remained high and low in the ICT. At the time of the breath release (i.e. phase III), the levels of O2Hb and HbT sharply decreased in the ECT. A reduction in the ICT was less visible, probably because brain saturation was already compromised. In the following seconds (i.e. phase IV), there was a sharp increase of O2Hb and HbT in the ECT and ICT, with a slight time-delay between the two. Having completed the VM, the parameters of O2Hb and HbT in the ECT and ICT gradually returned to the values of the resting state. Notice that significant changes in O2Hb and HbT, coupled with a rather stable HHb, are consistent with modulations in the degree of perfusion of the tissue investigated, driven by changes in the pressure gradient between the head and the body at a constant tissue oxygen consumption. This is in agreement with previous studies on VM (50).

To test the statistical significance of the results, the peak changes in O2Hb, HHb, and HbT during the breath-holding (BH) phases of the VM (i.e., phases I and II) were extracted. A repeated-measures analysis of variance (rmANOVA) was performed considering two within-subject factors (i.e. Condition: Rest and VM; and Compartment: ICT and ECT). A significant effect was obtained when considering O2Hb for VM vs Rest (F=6.04, df=1-21, p=0.023) and ICT vs. ECT (F=35.51, df=1-21, p=∼0), and when considering HbT for VM vs Rest (F=7.28, df=1-21, p=0.014) and ICT vs. ECT (F=28.19, df=1-21, p=∼0). Importantly a highly significant interaction between factors was obtained, indicating a synergic effect between condition and compartment for O2Hb (F=35.56, df=1-21, p=∼0) and HbT (F=28.42, df=1-21, p=∼0). When performing a post-hoc analysis, a significant increase in O2Hb and HbT was found in the ECT when comparing VM to Rest (O2Hb: t=5.10, df=21, p=4.7·10^-5^; HbT: t=4.78, df=21, p=9.9·10^-4^) whereas a significant decrease in O2Hb and HbT was obtained in the ICT (O2Hb: t=-3.85, df=21, p=9.2·10^-4^; HbT: t=-3.74, df=21, p=1.0·10^-3^).

Moreover, when comparing the ECT and the ICT during the VM, a significant difference was obtained for both O2Hb (t=5.96, df=21, p=6.4·10^-6^) and HbT (t=5.32, df=21, p=2.8·10^-5^).

## 4 Discussion

### 4.1 Co-registration

The developed approach yielded an average displacement error, expressed as RMSE, of 5.5 mm (Figure 5), which importantly is well below the spatial resolution limit of DOT of ∼15 mm (51). This means that the Artec Leo images, through the method presented here, can be applied to obtain an accurate co-registration of the optodes onto an anatomical template or a subject-specific anatomical image, with similar performance to electromagnetic digitization.

It is important to stress that the method reported here relies upon measured distances between the white dots and the optodes from a previously performed electromagnetic digitization. Hence, the developed approach for the co-registration of optical data on TBI patients in the ICU would require digitizing the white dots and the optical array on a phantom head or a healthy volunteer, with a head circumference similar to that of the subject that is going to be measured. Using a digitized helmet, the measurements performed on the TBI patient using an Artec Leo image would allow clinicians to correctly locate the array onto the patient’s CT scan or MRI, where the same fiducials can be selected.

Remarkably, this method can be performed without additional CT scans beyond the one performed upon admission for clinical assessment. The co-registration to the initial CT scan allows clinicians to prioritise the optical analysis of the most affected and clinically significant regions. The optical assessment of co-registered regions also allows clinicians to link the optical parameters and their functional evolution, to known lesions on the CT scans and their anatomical evolution. It should be mentioned that the ability to assess the optical properties of a brain lesion would depend on its size, so lesions smaller than the margin of error between the two co-registrations are likely to be overlooked, however, this problem is inherent to the limited spatial resolution of DOT (51). The decoupling of the commencement of DOT recordings from a targeted anatomical image would be useful either in the most severe cases of TBI, who for example can experience high ICP levels when supine or have in place extracorporeal systems (e.g., hemodialysis), all of which can be obstacles to performing further imaging, or in cases where clinicians are prioritising other patients and a second anatomical imaging may be postponed.

The degree of error between the two techniques may differ slightly from the 5.5 mm RMSE measured. The coordinates from Artec Leo can fail to exactly identify the digitized spots within the area of the white dots, so the absolute value of RMSE could be partially due to the testing analysis itself rather than the method implemented. However, this miscalculation can also affect the optical analysis, as the optode co-registration by Artec Leo is based on the measured distances between white dots and optodes by the electromagnetic digitization. The small sizes of the white dots mitigate, to a greater or less extent, this error. The holders’ width, especially for the detectors, can be another cause of inaccuracy in the optical analysis, because there could be a mismatch between the digitized spot within the holders and the real loci of the optodes.

Of note is that the analysis was performed on white dots that were uniformly distributed around the optode locations. Thus, it is essential for future helmets to maintain the uniformity of white dots in their design to accurately recover the optode locations on the anatomical image.

Although the results show a plateau of RMSE by using ten white dots, applications of this method on TBI for research purposes led us to think that helmets should possess as many white dots as can be fitted onto the exposed surface. This is because acquiring an optimal 3D image may not be always possible in hospital settings and blurred images may impair the retrieval of all the white dots. Moreover, the availability of white dots uniformly covering, not only the area around the optodes, but the whole head surface, may allow to employ more complex algorithms that may further reduce the optodes placement error (16, 17).

### 4.2 Optical Analysis

The fluctuations of O2Hb, HHb, and HbT levels in the ICT due to the VM were separated in the analysis from those in the ECT. In both layers, the results conformed with the expected changes during the different phases of the VM. This consistency validates the optical system as being capable of monitoring changes in brain Hb concentration through fNIRS-DOT analysis.

A decrease of cerebral saturation during VM was also reported in studies that used NIRS devices (27, 32, 34, 52–54). However, these analyses were limited to measurement of the decrease of cerebral saturation, and the ECT signal plausibly contaminated them by an unknown value. On the contrary, the fNIRS-DOT analysis reported herein allowed for a comparison of the changes of levels of Hb in the ICT and ECT, providing a better understanding of how the detected optical signal relates to brain physiology. Similarly to other studies, we were unable to quantify the role played by the ECT in the ICT signal, however the strong hemodynamic signal in the ICT leads us to think that no matter the extent of this contamination, it would be insufficient to obscure the tracking of the ICT signal during the VM (55). We believe that our test validated the aims of the study: (i) to test the ability of ICT monitoring, and (ii) to separate the ICT-ECT signals.

These results are in agreement with a pilot study which was performed without the extensive use of short channels, leading us to think that the co-registration process implemented in both studies is a critical component in these optical results (56). This is in agreement with other studies that showed that co-registering the optical signal with a structural image could improve the imaging resolution and the localisation accuracy (57, 58). Furthermore, Clancy et al. specifically reported an improvement in the accuracy of the signal detected during VM in simulated studies, by co-registering the optical data with a structural image (59, 60). However, in a study on a healthy volunteer, the co-registration of the optical signal from a high-density patch into an atlas did not yield the same capacity to retrieve physiological changes of tissue saturation associated with the VM as in the simulations (61). This limited capacity in an *in vivo* analysis may be explained to some extent by a suboptimal co-registration process compared with the co-registration system presented herein. Furthermore, as mentioned in the introduction, the co-registration of the optical signal with a subject-specific image may be particularly significant for the assumption about the baseline optical parameters in TBI patients to be valid. Layer thicknesses and optical properties can vary significantly between individuals and across different areas of the head (62, 63). These differences can be even more significant in TBI patients due to brain lesions (e.g., ischemia, hemorrhage) or surgical interventions (e.g., decompressive craniectomy) (11). The co-registration of the optical analysis into a subject-specific image takes into account the individual anatomy of these layers and brain lesions.

### 4.3 Limitations

#### 4.3.1 Absence of arterial blood pressure monitoring

Grading of the VM based on the changes in blood pressure was not possible. This may have affected the results as the quality of the performance of the VM may have differed across participants due to varying levels of commitment to the task and levels of cardiovascular fitness. Furthermore, it is not possible to perform the VM in the supine position at the same level as in the orthostatic one (26, 33). This further highlights the risk of different intensities of VM across participants if the VM is not performed with sufficient strength.

The lack of blood pressure monitoring also limited the separation of the different phases of the VM to an analysis of the expected changes of levels of Hb at different times without accurately correlating them to the blood pressure changes.

#### 4.3.2 Absence of subject-specific structural image

An MRI template was used in place of the subject-specific structural image for the co-registration of the optical data. This reduced the accuracy of the fitting procedure of the probes on the participants’ scalps due to their different head shapes. Moreover, the approximation also affected the optical data analysis, because the assumed optical properties of the different head layers were probably not anatomically identical among subjects (e.g., layer thickness).

#### 4.3.3 Global hemodynamic response during Valsalva manoeuvre

The cerebral perfusion and tissue saturation in TBI patients can vary significantly across the brain, while the VM triggers a similar hemodynamic response across the whole brain (64, 65). Therefore, the ability to separate the different hemodynamic across the brain of TBI patients could not be determined from the results obtained.

#### 4.3.4 Limited field of view

Although studies which investigated a broader coverage of the head using DOT have been presented for clinical applications in non-ICU scenarios (e.g., Parkinson syndrome), we limited the analysis to the frontal lobes to address the standard of care in ICU (44). In the future, new studies with a broader field of acquisition should be implemented, while maintaining the standard of care in acute TBI patients.

#### 4.3.5 Limited number of probes and wavelengths

Our study was limited to 15 detectors and 16 sources at two wavelengths (690 and 830 nm) mainly designed to measure O2Hb and HHb levels. Future studies should apply HD (high density) DOT to further improve the optical accuracy, as well as using different wavelengths to make the recording more suitable for ICG measurement (6).

#### 4.3.6 Absence of phase-shift analysis

Though a frequency-domain device was used, phase data were not implemented into the optical analysis. From results on simulated and phantom studies, analysis of the phase-shift can, theoretically, further increase imaging resolution and accuracy (66, 67). Future studies should explore this potentiality.

## 5 Conclusions

The results suggest that our method is capable of addressing the foreseeable problems associated with DOT neuromonitoring in TBI patients in the ICU. This procedure could enable fNIRS-DOT and contrast-enhanced DOT recordings as part of a multimodal monitoring on TBI patients upon hospital admission.

Future research should test other probe-position geometries up to HD-DOT, which may further increase the capability of the optical analysis to separate the signal from the different head layers and to monitor brain lesions. Clinical studies should be designed so that DOT measurements are included in a multimodal monitoring.

## 7 Code, Data, and Materials Availability

Code and anonymized optical data are available in Matlab format in FigShare at https://doi.org/10.6084/m9.figshare.25866682.v1

## Acknowledgments

This article presents independent research funded by the National Institute for Health Research Surgical Reconstruction and Microbiology Research Centre (NIHR SRMRC), partnership between University Hospitals Birmingham NHS Foundation Trust, the University of Birmingham, and the Royal Centre for Defence Medicine. The views expressed are those of the authors and not necessarily those of the NHS, the NIHR or the Department of Health. Dr Mario Forcione thanks Mr Michael Orgill from Gill Learning (https://www.gill-learning.co.uk) for his academic advice and proof reading.

## 8 Author Biographies and Photographs

Mario Forcione is a specialty registrar in the University of Milan-Bicocca in Anaesthesia and Intensive Care and has an honorary contract with the University Hospital Birmingham NHS Foundation Trust. In 2022, he received his Ph.D. from the University of Birmingham in clinical applications of diffuse optics to brain trauma. In 2015, he received his BS in Medicine from La

Sapienza, University of Rome. His work relates to the application of diffuse optics to the care of mild, moderate, and severe traumatic brain injury patients.

Antonio Maria Chiarelli is an Associate Professor in Applied Physics at the University of Chieti- Pescara. He received his BS and MS in Physics Engineering at the Polytechnic of Milan and his PhD in Neuroimaging at the University of Chieti. He worked as a Post Doctoral researcher at Beckman Institute, IL, USA. His research focuses on neuroimaging with Magnetic Resonance Imaging and Diffuse Optics for studying the human brain function and physiology.

David Perpetuini is an Assistant Professor in Biomedical Engineering at the University of Chieti- Pescara. He graduated in Biomedical Engineering from the Marche Polytechnic University of Ancona and he obtained his PhD in Neuroimaging at the University of Chieti. His research mainly concerns the study of brain and autonomic nervous system activity through functional near- infrared spectroscopy (fNIRS) and thermal imaging.

Guy Perkins completed his BSc in Physics in 2018 and then his PhD in 2024, both at the University of Birmingham. During his PhD, he investigated the development of FD-fNIRS and DOT for clinical applications. He is now a post-doctoral research fellow at the University of Padova, investigating preterm infants’ brain health in response to hyper/hypo glycemia.

Andrew Stevens is a Specialist Registrar in Neurosurgery, and academic researcher at the University of Birmingham in the fields of traumatic brain and spinal cord injury. He graduated from the University of Birmingham in 2016 with a Bachelors in Medicine and Surgery (MBChB), following completion of a BMedSc in 2013. He is currently completing a PhD in neurotrauma alongside his clinical practice.

Biographies and photographs for the other authors are not available.

## References

1. A. I. R. Maas et al., “Traumatic brain injury: integrated approaches to improve prevention, clinical care, and research,” Lancet Neurol 16(12), 987–1048 (2017).

2. D. J. Davies et al., “Near-Infrared Spectroscopy in the Monitoring of Adult Traumatic Brain Injury: A Review,” Journal of neurotrauma 32(13), 933–941 (2015).

3. M. S. Green, S. Sehgal, and R. Tariq, “Near-Infrared Spectroscopy: The New Must Have Tool in the Intensive Care Unit?,” Seminars in cardiothoracic and vascular anesthesia 20(3), 213–224 (2016).

4. I. K. Haitsma, and A. I. Maas, “Monitoring cerebral oxygenation in traumatic brain injury,” Progress in brain research 161(207-216 (2007).

5. M. Forcione et al., “Cerebral perfusion and blood–brain barrier assessment in brain trauma using contrast-enhanced near-infrared spectroscopy with indocyanine green: A review,” Journal of Cerebral Blood Flow & Metabolism 0271678X20921973 (2020).

6. M. Forcione et al., “Dynamic contrast-enhanced near-infrared spectroscopy using indocyanine green on moderate and severe traumatic brain injury: a prospective observational study,” Quantitative Imaging in Medicine and Surgery (2020).

7. M. Ganau et al., “Breakthrough in the assessment of cerebral perfusion and vascular permeability after brain trauma through the adoption of dynamic indocyanin green- enhanced near-infrared spectroscopy,” Quantitative Imaging in Medicine and Surgery 10(11), 2081–2084 (2020).

8. D. J. Davies et al., “Cerebral Oxygenation in Traumatic Brain Injury: Can a Non-Invasive Frequency Domain Near-Infrared Spectroscopy Device Detect Changes in Brain Tissue Oxygen Tension as Well as the Established Invasive Monitor?,” Journal of neurotrauma 36(7), 1175–1183 (2019).

9. S. R. Leal-Noval et al., “Invasive and noninvasive assessment of cerebral oxygenation in patients with severe traumatic brain injury,” Intensive Care Med 36(8), 1309–1317 (2010).

10. W. Weigl et al., “Application of optical methods in the monitoring of traumatic brain injury: A review,” J Cereb Blood Flow Metab 36(11), 1825–1843 (2016).

11. M. Forcione et al., “Mismatch between Tissue Partial Oxygen Pressure and Near-Infrared Spectroscopy Neuromonitoring of Tissue Respiration in Acute Brain Trauma: The Rationale for Implementing a Multimodal Monitoring Strategy,” International Journal of Molecular Sciences 22(3), 1122 (2021).

12. I. J. Bigio, and S. Fantini, Quantitative Biomedical Optics: Theory, Methods, and Applications, Cambridge University Press, Cambridge (2016).

13. M. D. Wheelock, J. P. Culver, and A. T. Eggebrecht, “High-density diffuse optical tomography for imaging human brain function,” Review of Scientific Instruments 90(5), 051101 (2019).

14. T. Austin et al., “Three dimensional optical imaging of blood volume and oxygenation in the neonatal brain,” NeuroImage 31(4), 1426–1433 (2006).

15. G. Giacalone et al., “Time-domain near-infrared spectroscopy in acute ischemic stroke patients,” Neurophotonics 6(1), 015003 (2019).

16. C. Whalen et al., “Validation of a method for coregistering scalp recording locations with 3D structural MR images,” Hum Brain Mapp 29(11), 1288–1301 (2008).

17. A. M. Chiarelli et al., “Comparison of procedures for co-registering scalp-recording locations to anatomical magnetic resonance images,” Journal of biomedical optics 20(1), 016009 (2015).

18. 18. E. E. Moore, D. V. Feliciano, and K. L. Mattox, Trauma, Eighth Edition, McGraw-Hill Education (2017).

19. 19. C. H. Tan, et al., “Mapping cerebral pulse pressure and arterial compliance over the adult lifespan with optical imaging,” PLoS One 12(2), e0171305 (2017).

20. L. He et al., “Noninvasive continuous optical monitoring of absolute cerebral blood flow in critically ill adults,” Neurophotonics 5(4), 045006 (2018).

21. H. Dehghani et al., “Near infrared optical tomography using NIRFAST: Algorithm for numerical model and image reconstruction,” Communications in numerical methods in engineering 25(6), 711–732 (2008).

22. K. E. Mathewson et al., “Dynamics of Alpha Control: Preparatory Suppression of Posterior Alpha Oscillations by Frontal Modulators Revealed with Combined EEG and Event-related Optical Signal,” Journal of cognitive neuroscience 26(10), 2400–2415 (2014).

23. C. S. Robertson et al., “Clinical evaluation of a portable near-infrared device for detection of traumatic intracranial hematomas,” Journal of neurotrauma 27(9), 1597–1604 (2010).

24. M. Forcione et al., “Tomographic Task-Related Functional Near-Infrared Spectroscopy in Acute Sport-Related Concussion: An Observational Case Study,” Int J Mol Sci 21(17), (2020).

25. F. P. Tiecks et al., “Effects of the valsalva maneuver on cerebral circulation in healthy adults. A transcranial Doppler Study,” Stroke 26(8), 1386–1392 (1995).

26. F. Pott et al., “Middle cerebral artery blood velocity during a valsalva maneuver in the standing position,” Journal of applied physiology (Bethesda, Md. : 1985) 88(5), 1545–1550 (2000).

27. B. G. Perry et al., “Cerebral hemodynamics during graded Valsalva maneuvers,” Frontiers in physiology 5(349 (2014).

28. M. H. Wilson, “Monro-Kellie 2.0: The dynamic vascular and venous pathophysiological components of intracranial pressure,” J Cereb Blood Flow Metab 36(8), 1338–1350 (2016).

29. H. Prabhakar et al., “Intracranial pressure changes during Valsalva manoeuvre in patients undergoing a neuroendoscopic procedure,” Minim Invasive Neurosurg 50(2), 98–101 (2007).

30. M. J. Haykowsky et al., “Resistance exercise, the Valsalva maneuver, and cerebrovascular transmural pressure,” Med Sci Sports Exerc 35(1), 65–68 (2003).

31. J. C. Greenfield, Jr., J. C. Rembert, and G. T. Tindall, “Transient changes in cerebral vascular resistance during the Valsalva maneuver in man,” Stroke 15(1), 76–79 (1984).

32. D. Davies et al., “Comparison of near infrared spectroscopy with functional MRI for detection of physiological changes in the brain independent of superficial tissue,” The Lancet 387(S34 (2016).

33. B. G. Perry et al., “The cerebrovascular response to graded Valsalva maneuvers while standing,” Physiol Rep 2(2), e00233 (2014).

34. D. J. Davies et al., “The Valsalva maneuver: an indispensable physiological tool to differentiate intra versus extracranial near-infrared signal,” Biomed. Opt. Express 11(4), 1712–1724 (2020).

35. J. Steinbrink et al., “Determining changes in NIR absorption using a layered model of the human head,” Physics in Medicine and Biology 46(3), 879–896 (2001).

36. D. J. Davies, “Cerebral near infra-red spectroscopy in traumatic brain injury as a potential independent monitoring modality and alternative to invasive tissue oxygen tension sensors,” in *School of Clinical and Experimental Medicine* University of Birmingham, University of Birmingham UBIRA E THESIS (2017).

37. K. J. Lee et al., “Non-invasive detection of intracranial hypertension using a simplified intracranial hemo- and hydro-dynamics model,” BioMedical Engineering OnLine 14(1), 51 (2015).

38. S. R. Mousavi et al., “Measurement of in vivo cerebral volumetric strain induced by the Valsalva maneuver,” J Biomech 47(7), 1652–1657 (2014).

39. B. B. Ertl-Wagner et al., “Demonstration of periventricular brain motion during a Valsalva maneuver: description of technique, evaluation in healthy volunteers and first results in hydrocephalic patients,” European Radiology 11(10), 1998–2003 (2001).

40. A. M. Chiarelli et al., “A kurtosis-based wavelet algorithm for motion artifact correction of fNIRS data,” NeuroImage 112(128-137 (2015).

41. D. Perpetuini et al., “A Motion Artifact Correction Procedure for fNIRS Signals Based on Wavelet Transform and Infrared Thermography Video Tracking,” Sensors 21(15), 5117 (2021).

42. A. Ishimaru, “Diffusion of light in turbid material,” Appl. Opt. 28(12), 2210–2215 (1989).

43. K. D. Paulsen, and H. Jiang, “Spatially varying optical property reconstruction using a finite element diffusion equation approximation,” Medical physics 22(6), 691–701 (1995).

44. A. T. Eggebrecht et al., “Mapping distributed brain function and networks with diffuse optical tomography,” Nat Photonics 8(6), 448–454 (2014).

45. F. Qianqian, and D. A. Boas, “Tetrahedral mesh generation from volumetric binary and grayscale images,” 2009 IEEE International Symposium on Biomedical Imaging: From Nano to Macro 1142-1145 (2009).

46. K. J. Friston et al., “Statistical parametric maps in functional imaging: A general linear approach,” Human Brain Mapping 2(4), 189–210 (1994).

47. W. Penny, et al., Statistical Parametric Mapping: The Analysis of Functional Brain Images, Elsevier: Amsterdam, The Netherlands (2011).

48. F. Tian, and H. Liu, “Depth-compensated diffuse optical tomography enhanced by general linear model analysis and an anatomical atlas of human head,” NeuroImage 85 **Pt 1**(166- 180 (2014).

49. W. G. Zijlstra, A. Buursma, and W. P. Meeuwsen-van der Roest, “Absorption spectra of human fetal and adult oxyhemoglobin, de-oxyhemoglobin, carboxyhemoglobin, and methemoglobin,” Clin Chem 37(9), 1633–1638 (1991).

50. A. Bluestone et al., “Three-dimensional optical tomography of hemodynamics in the human head,” Opt Express 9(6), 272–286 (2001).

51. A. M. Chiarelli et al., “Combining energy and Laplacian regularization to accurately retrieve the depth of brain activity of diffuse optical tomographic data,” Journal of biomedical optics 21(3), 36008 (2016).

52. 52. M. Clancy, et al., “Comparison of Neurological NIRS signals during standing Valsalva maneuvers, pre and post vasopressor injection,” in Diffuse Optical Imaging V H. Dehghani, and P. Taroni, Eds., Spie-Int Soc Optical Engineering, Bellingham (2015).

53. D. Canova et al., “Inconsistent detection of changes in cerebral blood volume by near infrared spectroscopy in standard clinical tests,” Journal of applied physiology (Bethesda, Md. : 1985) 110(6), 1646–1655 (2011).

54. R. Saager, and A. Berger, “Measurement of layer-like hemodynamic trends in scalp and cortex: implications for physiological baseline suppression in functional near-infrared spectroscopy,” Journal of biomedical optics 13(3), 034017 (2008).

55. N. Eleveld et al., “The Influence of Extracerebral Tissue on Continuous Wave Near-Infrared Spectroscopy in Adults: A Systematic Review of In Vivo Studies,” J Clin Med 12(8), (2023).

56. 56. M. Forcione, “Neuromonitoring in mild, moderate, and severe acute brain trauma using non-invasive diffuse optics,” in College of Medical and Dental Sciences, p. 230, University of Birmingham (2021).

57. D. A. Boas, and A. M. Dale, “Simulation study of magnetic resonance imaging-guided cortically constrained diffuse optical tomography of human brain function,” Appl Opt 44(10), 1957–1968 (2005).

58. D. A. Boas, A. M. Dale, and M. A. Franceschini, “Diffuse optical imaging of brain activation: approaches to optimizing image sensitivity, resolution, and accuracy,” NeuroImage 23 **Suppl 1**(S275-288 (2004).

59. M. Clancy et al., “Monitoring the Injured Brain - Registered, patient specific atlas models to improve accuracy of recovered brain saturation values,” in Diffuse Optical Imaging V H. Dehghani, and P. Taroni, Eds., Spie-Int Soc Optical Engineering, Bellingham (2015).

60. M. Clancy et al., “Improving the quantitative accuracy of cerebral oxygen saturation in monitoring the injured brain using atlas based Near Infrared Spectroscopy models,” Journal of Biophotonics 9(8), 812–826 (2016).

61. 61. M. Clancy, “Application and development of high-density functional near infrared spectroscopy for traumatic brain injury,” in School of Chemistry, University of Birmingham, University of Birmingham UBIRA E THESES (2017).

62. L. Gagnon et al., “Short separation channel location impacts the performance of short channel regression in NIRS,” NeuroImage 59(3), 2518–2528 (2012).

63. F. Scholkmann et al., “Absolute Values of Optical Properties (mua, mus, mueff and DPF) of Human Head Tissue: Dependence on Head Region and Individual,” Adv Exp Med Biol 1072(325-330 (2018).

64. J. P. Coles et al., “Effect of hyperventilation on cerebral blood flow in traumatic head injury: clinical relevance and monitoring correlates,” Crit Care Med 30(9), 1950–1959 (2002).

65. W. B. Baker et al., “Continuous non-invasive optical monitoring of cerebral blood flow and oxidative metabolism after acute brain injury,” Journal of Cerebral Blood Flow & Metabolism 39(8), 1469–1485 (2019).

66. G. A. Perkins, A. T. Eggebrecht, and H. Dehghani, “Quantitative evaluation of frequency domain measurements in high density diffuse optical tomography,” Journal of biomedical optics 26(5), (2021).

67. G. A. Perkins, A. Eggebrecht, and H. Dehghani, “Multi-Modulated frequency domain high densitydiffuse optical tomography,” Biomed. Opt. Express 13((2022).

